# A simulation-based method to inform serosurvey designs for estimating dengue force of infection using existing blood samples

**DOI:** 10.1101/2023.04.07.23288282

**Authors:** Anna Vicco, Clare McCormack, Belen Pedrique, John H. Amuasi, Anthony Afum-Adjei Awuah, Christian Obirikorang, Nicole S. Struck, Eva Lorenz, Jürgen May, Isabela Ribeiro, Gathsaurie Neelika Malavige, Christl A. Donnelly, Ilaria Dorigatti

## Abstract

The extent to which dengue virus has been circulating in Africa is largely unknown. Testing available blood samples from previous cross-sectional serological surveys offers a convenient strategy to investigate past dengue infections, as such serosurveys provide the ideal data to reconstruct the age-dependent immunity profile of the population and to estimate the average per-capita annual risk of infection; the force of infection (FOI), which is a fundamental measure of transmission intensity.

In this study, we present a novel methodological approach to inform the size and age distribution of blood samples to test when samples are acquired from previous surveys. The method was used to inform a dengue seroprevalence survey which is currently being conducted in Ghana by the Drug for Neglected disease initiative, utilizing samples previously collected for a SARS-CoV-2 serosurvey.

The method described in this paper can be employed to determine sample sizes and testing strategies for different diseases and transmission settings.

**Author summary:** The historical circulation of dengue virus in Africa is still poorly understood, and age-stratified seroprevalence surveys can provide the data to quantify population exposure to dengue and its transmission intensity.

In this work, we developed a simulation-based method that can be used to identify the sample sizes and age-distribution of the samples needed to obtain informative estimates of dengue force of infection from existing blood samples. We apply this method to data obtained from a SARS-CoV-2 serological survey, previously conducted in three cities in Ghana.

The proposed method can be used to design serological surveys for other pathogens when using existing blood samples with accompanying age and location are available.

## Introduction

Dengue virus (DENV) is one of the most rapidly spreading vector-borne diseases worldwide and is a burden on global public health [1]. In the last 50 years, the incidence of dengue infection has grown more than 30-fold [2], with recent estimates suggesting that up to 105 million people are infected each year [3]. Currently, there are four antigenically distinct dengue serotypes (DENV1-4), which can co-circulate [2]. Infection with one serotype confers lifelong protection against the same serotype and temporary cross-protection against heterologous serotypes [1]. There is epidemiological evidence that secondary infections are associated with more severe disease [4], due to an immuno-pathological phenomenon known as antibody dependent enhancement (ADE) [5].

The severity of a dengue infection can range from a mild fever to life-threatening complications, such as fever progressing to haemorrhage, dengue shock syndrome and organ failure, particularly in children [6].

According to the World Health Organisation (WHO), mortality rates in severe dengue cases range from 1% to 20% [1], and whilst the differential strengths of health care systems certainly play a role, there are still fundamental gaps in our understanding of the individual - and population-level risk factors leading to hospitalization and death [7]. Currently, there is no treatment for dengue, but new antiviral drugs, including a pan-serotype dengue virus inhibitor, which has demonstrated a high efficacy in mouse models, have entered clinical testing [8].

The first licensed dengue vaccine, Dengvaxia (CYD-TDV) developed by Sanofi Pasteur, is only recommended for use in subjects with a previously confirmed dengue infection [9]. A new dengue live attenuated vaccine TAK-003, developed by Takeda, has been recently licensed in several countries [10] and has demonstrated a cumulative efficacy of more than 70% against virologically confirmed dengue disease in the two years following vaccination, with a slight decline in protection during the second year [11], differences by serotype [12] and an efficacy against hospitalization of more than 77% three years post vaccination [13]. Another live attenuated tetravalent dengue vaccine, developed by the Butanan Institute in collaboration with the US National Institutes of Health (NIH), has recently finished a phase II clinical trial where it showed seroconversion in 77% to 92% of vaccinated subjects, with variations observed between dengue-naïve and pre-exposed participants at vaccination [14]. Moreover, novel biological vector control strategies, such as the release of Wolbachia-carrying mosquitoes, have been evaluated in cluster-randomized trials across multiple sites in endemic countries [15], demonstrating an efficacy of over 77% on average against virologically confirmed dengue in a cluster-randomized trial in Indonesia [16].

With ongoing changes in climate, demography, socioeconomic structure and growing urbanization [17], there are concerns that the geographical range of dengue circulation will expand in the coming decades. Moreover, estimates suggest it may affect Sub-Saharan Africa in particular [18–20], a region where there is great uncertainty around the historical circulation of dengue and the extent of exposure among populations. Despite evidence that all four dengue serotypes are present in Africa, mainly in the east of the continent [21], dengue surveillance is limited, with cases often only reported during outbreaks, in observational studies and in returning travellers [22]. The absence of systematic mosquito surveillance, along with insufficient resources to build training and testing capacity, have been identified as key gaps for the development of arbovirus surveillance and disease control across the African region [23]. Furthermore, the documented invasion of *Aedes albopictus* across Africa [20] provides yet another reason for strengthening disease surveillance across countries.

Due to high rates of asymptomatic infection [17,24] and misclassification [2], the burden of dengue in Africa is still poorly understood and largely underestimated [25,26]. Serological surveys are the gold standard for understanding the historical transmission of dengue across settings [27]. However, only 17 age-stratified seroprevalence surveys have been conducted in Africa up to now [3]. Cross-sectional serological surveys provide the data to reconstruct the immunity profile of the population, i.e., the extent of dengue exposure as a function of age, which in turn can be used to estimate the force of infection (FOI), the per capita rate at which susceptible individuals become infected [28].

While the implementation of new seroprevalence surveys provides the opportunity to target optimal age groups to estimate the FOI, it also requires extensive resources, including time and operational capacity. However, these requirements can be reduced if it is possible to test blood samples collected in the context of other surveys. In this study, we aimed to develop a new method to inform the optimal testing strategy that uses existing blood samples with accompanying information on the age and residence location to estimate the FOI. While we focus on dengue, this method can be adopted to estimate the FOI for other infectious pathogens across transmission settings.

## Materials and methods

### Blood samples

The blood samples available for testing were collected in a SARS-CoV-2 seroprevalence study conducted in three cities in Ghana (Accra, Kumasi and Tamale) between February 2021 and February 2022 by Bernard Nocht Institute for Tropical Medicine (BNITM) and Kumasi Centre for Collaborative Research in Tropical Medicine (KCCR) [29]. The available blood samples included 2,051 samples from participants aged 10 to 94 years; all samples were stored in a biobank at -80°C.

### Potential sampling scenarios

We simulated alternative sampling scenarios (Table 1) to select the sample sizes and their distribution. We denoted the scenario that included testing all available samples as ‘scenario 0’ and explored the following alternative scenarios: testing an equal number of samples in each age-group, using the minimum number of available samples by age-group (scenario A); testing all available samples in the younger age-groups (i.e., the first half of the age-groups), and half of the minimum number of samples across the young-age groups for the older age-groups (scenario B); testing all available samples for the older age-groups and half of the minimum number of samples across the old-age groups for the younger age-groups (scenario C); testing all available samples for the younger age-groups and half of the available number of samples for the older age-groups (scenario D) and testing all available samples for the older age-groups and half of the available number of samples for the younger age-groups (scenario E).

**Table 1.** Sample sizes for 5-year age categorization used in scenarios 0 to E in the three cities in Ghana.

| Age group<br>5-years | Accra |  |  |  |  |  | Kumasi |  |  |  |  |  | Tamale |  |  |  |  |  |
| --- | --- | --- | --- | --- | --- | --- | --- | --- | --- | --- | --- | --- | --- | --- | --- | --- | --- | --- |
|  | 0 | A | B | C | D | E | 0 | A | B | C | D | E | 0 | A | B | C | D | E |
| 10-14 | 20 | 20 | 20 | 16 | 20 | 10 | 22 | 22 | 22 | 17 | 22 | 11 | 14 | 13 | 14 | 6 | 14 | 7 |
| 15-19 | 57 | 20 | 57 | 16 | 57 | 28 | 58 | 22 | 58 | 17 | 58 | 29 | 64 | 13 | 64 | 6 | 64 | 32 |
| 20-24 | 81 | 20 | 81 | 16 | 81 | 40 | 88 | 22 | 88 | 17 | 88 | 44 | 100 | 13 | 100 | 6 | 100 | 50 |
| 25-29 | 90 | 20 | 90 | 16 | 90 | 45 | 82 | 22 | 82 | 17 | 82 | 41 | 113 | 13 | 113 | 6 | 113 | 56 |
| 30-34 | 69 | 20 | 69 | 16 | 69 | 34 | 65 | 22 | 65 | 17 | 65 | 32 | 118 | 13 | 118 | 6 | 118 | 59 |
| 35-39 | 64 | 20 | 64 | 16 | 64 | 32 | 50 | 22 | 50 | 17 | 50 | 25 | 73 | 13 | 73 | 6 | 73 | 36 |
| 40-44 | 44 | 20 | 10 | 44 | 22 | 44 | 61 | 22 | 11 | 61 | 30 | 61 | 76 | 13 | 7 | 76 | 38 | 76 |
| 45-49 | 66 | 20 | 10 | 66 | 33 | 66 | 65 | 22 | 11 | 65 | 32 | 65 | 32 | 13 | 7 | 32 | 16 | 32 |
| 50-54 | 45 | 20 | 10 | 45 | 22 | 45 | 51 | 22 | 11 | 51 | 26 | 51 | 41 | 13 | 7 | 41 | 20 | 41 |
| 55-59 | 38 | 20 | 10 | 38 | 19 | 38 | 34 | 22 | 11 | 34 | 17 | 34 | 13 | 13 | 7 | 13 | 6 | 13 |
| 60-64 | 31 | 20 | 10 | 31 | 16 | 31 | 43 | 22 | 11 | 43 | 22 | 43 | 23 | 13 | 7 | 23 | 12 | 23 |
| 65+ | 62 | 20 | 10 | 62 | 31 | 62 | 59 | 22 | 11 | 59 | 30 | 59 | 39 | 13 | 7 | 39 | 20 | 39 |
| **Total** | **667** | **240** | **441** | **382** | **524** | **475** | **678** | **264** | **431** | **415** | **522** | **495** | **706** | **156** | **524** | **260** | **594** | **464** |

### Simulation study & model fitting

We assumed that the average yearly FOI had been constant in time and across all age-groups, and that antibodies do not wane. Using these assumptions, the age-dependent seroprevalence (i.e., the probability of developing measurable antibodies upon exposure) could be modelled as detailed in Eq. 1, where *a*_*i*_ is the midpoint of age group *i, λ* corresponds to the average yearly FOI per serotype, based on the estimates obtained from Cattarino et al. [3], and *n* is the number of serotypes assumed to be circulating. For Ghana, we assumed *n* = 2 independently transmitting serotypes, following Bonney et al. [30].

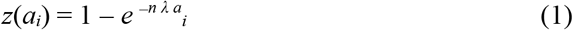

We assumed that the number of subjects exposed to dengue in each age-group was binomially distributed as a function of the number of samples tested in age group *i*, as described in Eq. 2

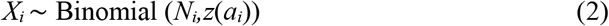

where *X*_*i*_ denotes the number of people testing seropositive in group *i, N*_*i*_ denotes the number of samples tested in age-group *i*, and *z*(*a*_*i*_) is the probability of testing seropositive in that age-group.

For each sampling scenario we denoted the age-specific sample sizes *N*_1_,*…,N*_*m*_ in age-groups 1,…,*m*, and simulated the expected age-stratified seroprevalence as shown in Eq. 1. For each scenario, we generated 1,000 datasets by sampling the number of seropositive samples *I*_1_,…,*I*_*m*_ from a binomial distribution with parameters *N*_1_,…,*N*_*m*_ and *z*_1_,…,*z*_*m*_, where *z* derives from Eq. 1, assuming the FOI estimates from Cattarino et al. [3] (Table S9). To account for sensitivity (*se)* and specificity (*sp)* of the test respectively, we simulated the number of samples testing positive *P*_1_,…, *P*_*m*_ among the *I*_1_,…,*I*_*m*_ samples as the sum of the true positive *TP*_1_,…*TP*_*m*_ and false positive *FP*_1_,…,*FP*_*m*_ samples, which were sampled from a binomial distribution as described in Eq. 3 - Eq. 5.

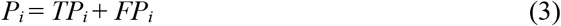

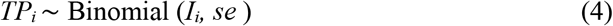

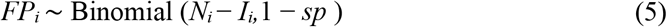

We assumed *se* = 0.892 and *sp* = 0.988 [32] and for each simulated dataset we fitted the catalytic model separately, thus obtaining 1,000 FOI estimates, denoted *λ*^∼^.

Using a binomial likelihood (Eq. 6) we reconstructed the posterior distribution of the FOI *λ* using the Hamiltonian Monte Carlo (HMC) algorithm coded in *CmdStanR* [31], having assumed a uniform prior distribution between zero and one, and sampling 3 chains of 5,000 iterations each, with a warming-up step of 1,000 iterations. Convergence was assessed visually and using the Rhat and effective sample size [31]. In the results we report the median and 95% credible interval of the posterior distribution of the FOI.

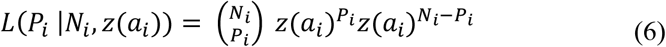

### Performance metrics and selection criteria

Three performance indexes were used to compare the FOI estimates *λ*^∼^ with the FOI used to generate the simulated data. Specifically, we calculated the median of the FOI under each scenario and age category, and then calculated the:

- **Bias**: the absolute value of the difference between the estimated FOI *λ*^∼^ and the *λ* used to simulate the data.
- **Uncertainty**: the width of the 95% credible interval (CrI) of the estimated FOI *λ*^∼^.
- **Coverage**: the number of times the FOI *λ* used to simulate the data falls within 95% CrI of the estimated FOI *λ*^∼^, across the 1,000 simulations.

We selected the optimal sample sizes and age-distributions based on the following criteria. Firstly, we retained only the scenarios with an upper bound of the 95% CrI of the bias within a 15% tolerance above the one obtained with scenario 0. Then, among these scenarios, we retained those with a coverage within 5% of the maximum coverage value across the selected scenarios. Finally, we selected the scenario with the smallest number of samples. We also compared *a posteriori* the uncertainty of the optimal scenario with the one of scenario 0 to check that the selected scenario did not introduce further uncertainty around the estimate.

In a sensitivity analysis, we ran the simulation study using 5 - and 10-year age categories to increase the sample sizes in each age group and explore the accuracy of the FOI estimates when using wider age groups.

## Results

A total of 2,051 blood samples from a previous seroprevalence study conducted in three cities in Ghana, Accra, Kumasi and Tamale [29], were available for testing for dengue antibodies (Table 1, scenario 0). Younger age-groups were less well represented in the study population than in the general population, with 24.6% of the participants belonging to the 10-24 years age group, compared to an expected 41.2% from the national age structure published in the 2022 Revision of World Population Prospects (WPP) (Fig 1). The 20-34 years age group was the most represented (39.3% of participants) [29].

**Fig 1.**
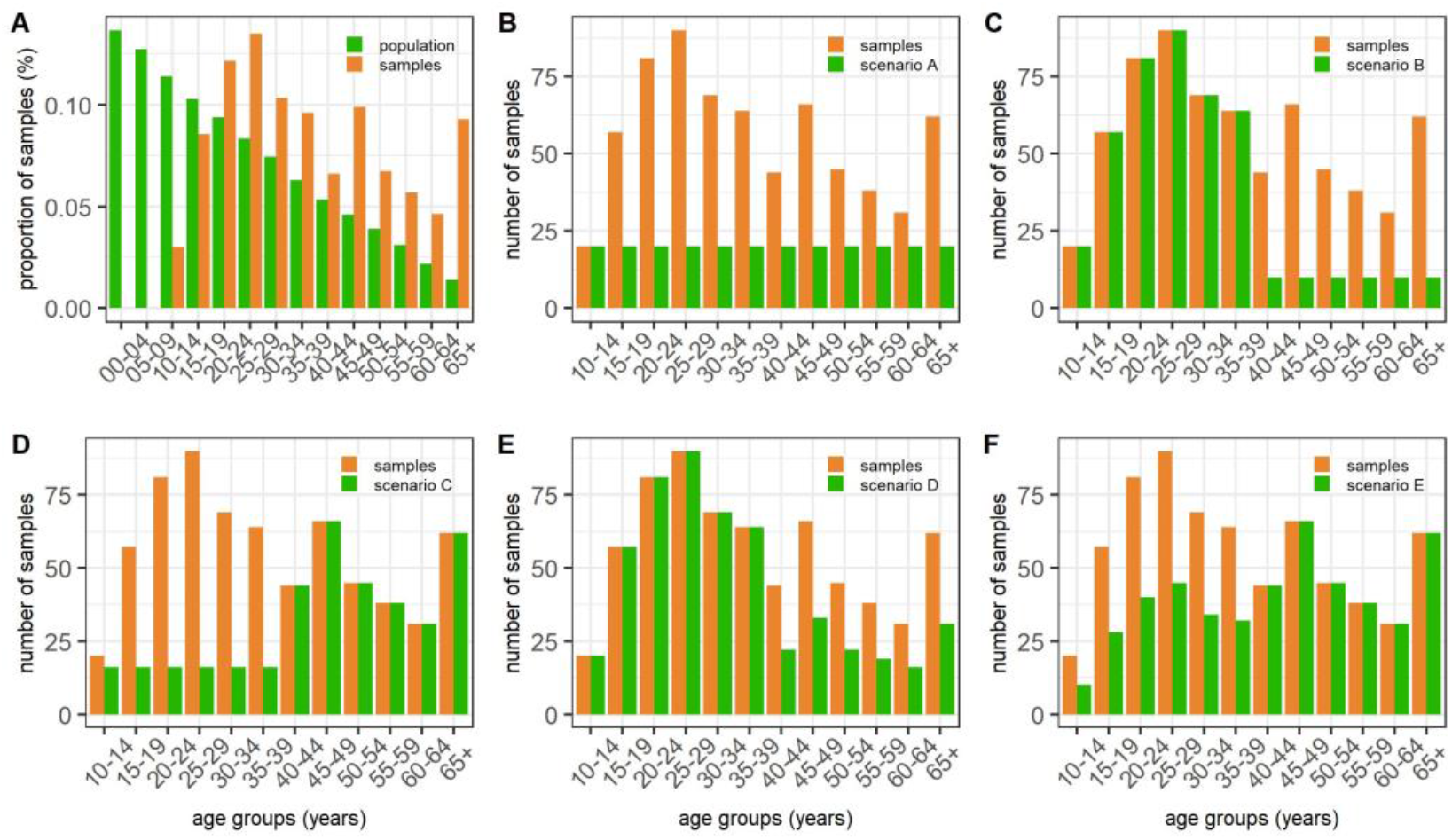
Blood sample distribution across several scenarios for the city of Accra using the 5-year age category. Panel A shows the distribution of the available samples across the age groups and compares them with the age-stratified population of Ghana (World Population Prospect 2021: https://population.un.org/wpp/). Panels B to F illustrate the sample distribution under each scenario (from A to E) compared to the baseline scenario 0 (which in the panel is indicated as “samples”).

Fig 2 shows the bias, uncertainty, and coverage obtained across all scenarios using the 5-year age categories for the three Ghanian cities. The corresponding figure for the 10-year age categories is presented in the Supplementary Material (Figure 1S).

**Fig 2.**
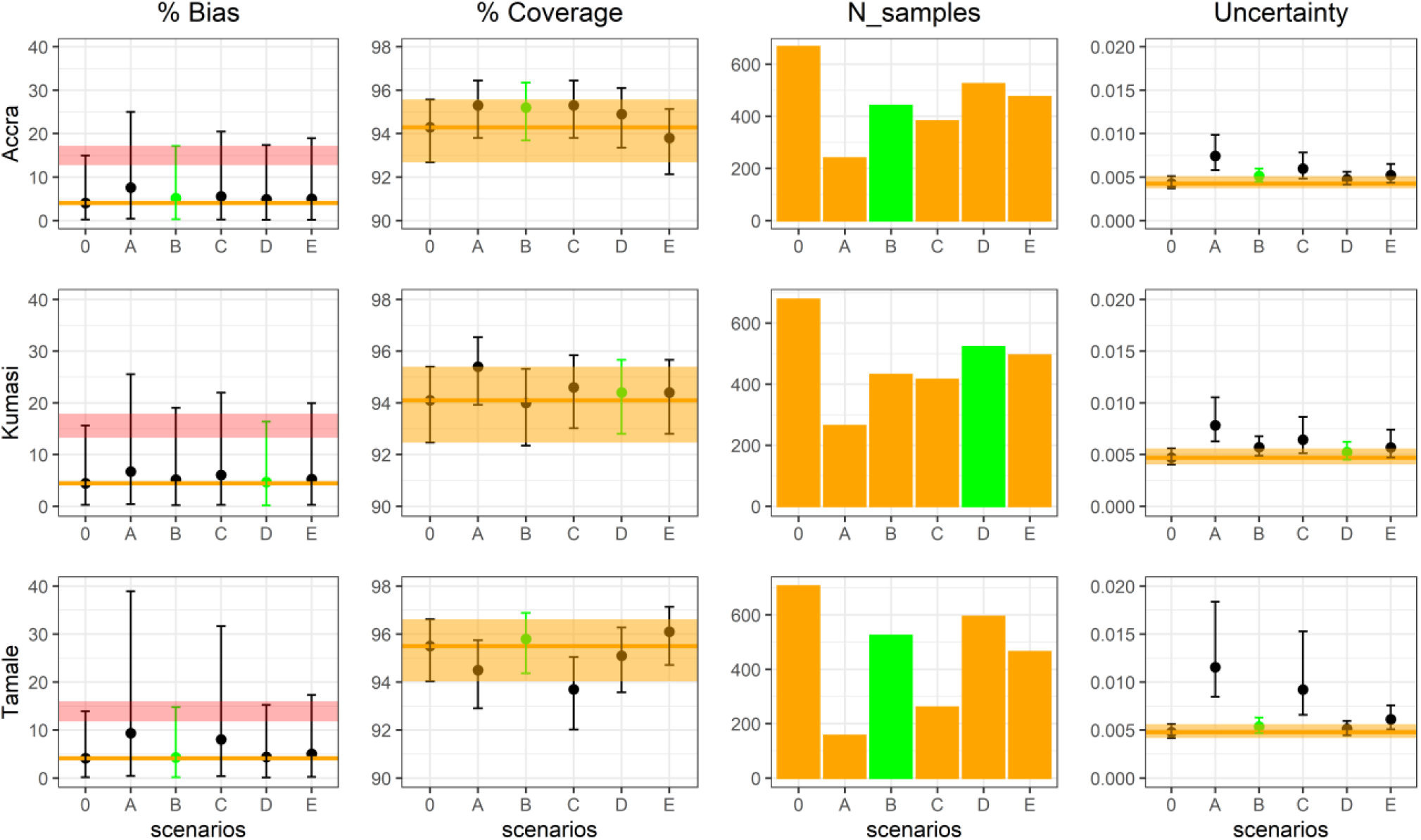
Summary of the accuracy metrics obtained for Accra, Kumasi and Tamale with 5-year age category across scenarios. The four columns represent respectively the bias, coverage, number of tested samples and uncertainty obtained for each scenario. The scenario highlighted in green indicates the selected scenario. The median bias and uncertainty are reported with their 95% CrI (columns 1 and 4), while the median coverage is reported with its 95% exact binomial CI. The orange line represents the median (columns 1, 2 and 4)and the orange ribbon represents the 95% CrI of the baseline scenario 0 (columns 2 and 4). The pink ribbon in the first column represents the 15% tolerance around the upper bound of the 95% CrI of scenario 0, which was used in the first step of the selection criterion.

Overall, we found that reducing the sample sizes did not bias the FOI estimates, except for scenario A (which had the smallest number of samples) where the central estimate was consistently larger. On the other hand, as expected, we found that reducing the sample sizes increased the uncertainty of the FOI estimates, depending on the number and distribution of the samples by age. On average, we observed a coverage above 90% (between 93% and 96%) across all analysed scenarios, with some variability depending on the city. Fig 2 shows that reducing the number of tests in certain age groups (such as in scenario B and highlighted in green) can increase the coverage without biasing the estimates.

In Ghana, using the FOI estimates from Cattarino et al. [3], we found that scenarios B and D, which included fewer samples from the older age-groups, showed a higher coverage and a lower uncertainty compared to the baseline scenario (scenario 0). This indicates that selecting the appropriate age distribution of the samples can help improve the accuracy of the FOI estimates, while at the same time reducing the sample sizes. Scenarios B and D provided the best combination of coverage, bias and uncertainty in both the 5-year and 10-year age categories for the three cities. In these scenarios, the distribution of samples in the two age categories led to a similar uncertainty, but the coverage was generally higher in the 5-year age category, which also required testing fewer samples than the 10-year group (a total of 1,487 instead of 1,610).

Table 2 shows the age group sample sizes for the selected scenarios and compares the total number of samples suggested using the method described in this paper to the initial number of available samples. Using the proposed selection criterion, the total sample size was reduced to 1,487 samples, rather than the available 2,051.

**Table 2.** Total number of samples in Ghana according to the selected scenario compared to all available samples.

| Age group | Accra | Kumasi | Tamale |
| --- | --- | --- | --- |
| Scenario | B | D | B |
| Total available | 667 | 678 | 706 |
| Total selected | 441 | 522 | 524 |

Fig 3 shows the expected seroprevalence profile and model fit obtained from the baseline scenario (scenario 0) versus those obtained from the selected scenario, indicating no significant deviation in model fit caused by the reduced samples sizes and adopted sampling strategy.

**Fig 3.**
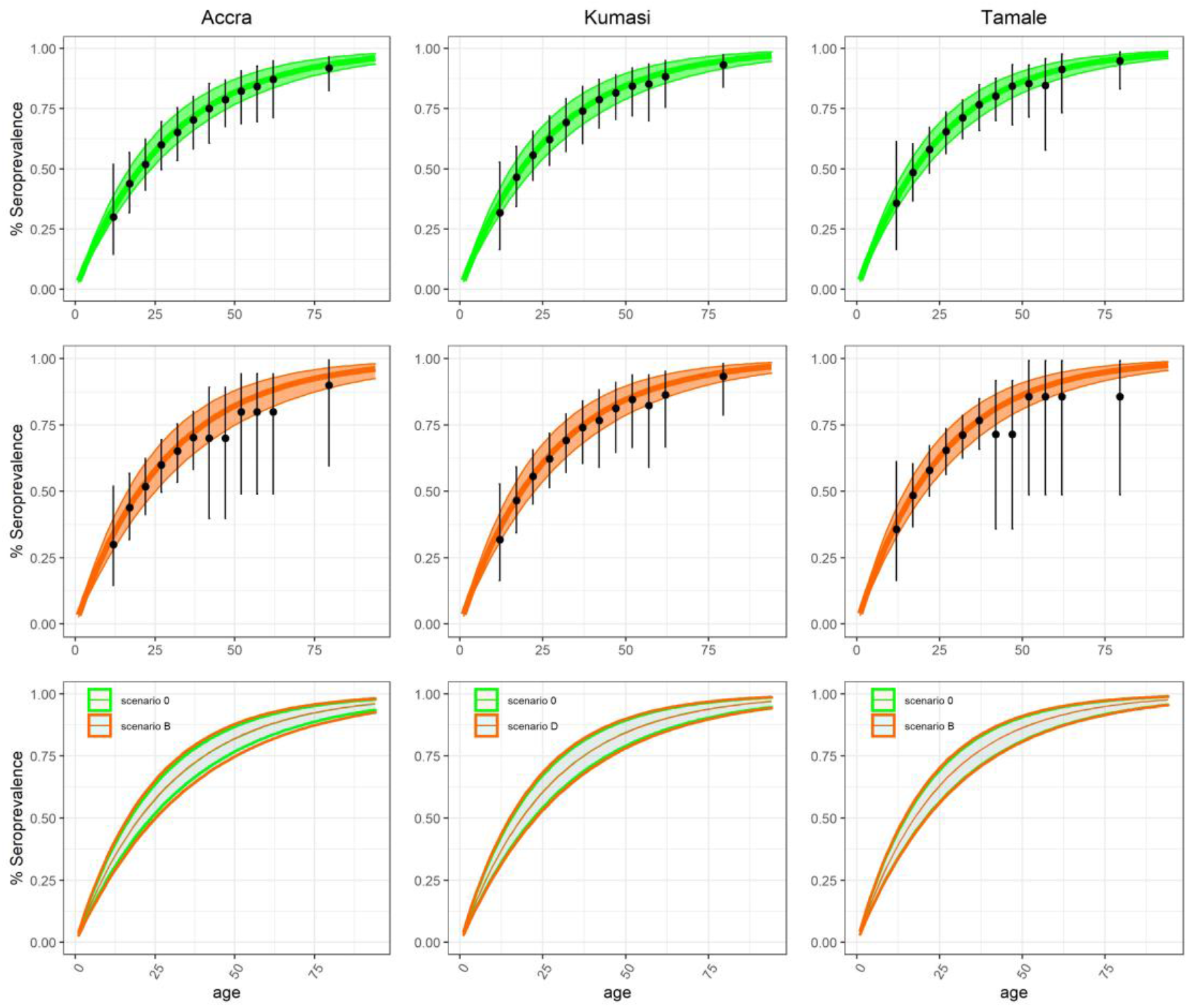
Model fit for the three cities under the 5-year age categorization. The first row represents the fit for scenario 0, where the median and 95% credible interval of the estimated seroprevalence are reported in green as a line and shaded area. The second row illustrates the fit under the selected scenario, optimal for each city, and is represented in orange. In both panels, the datasets that feed into each model are reported in black, with the error bar corresponding to the exact binomial confidence interval. In the third row we can compare the estimated seroprevalence in scenario 0 (in green shaded) and the optimal scenario (in orange) where the lines represent the 95% CrI.

## Discussion

Estimating the transmission intensity and burden of dengue globally is key to quantifying the current and future risk of infection and disease, and health care demand. In addition, it is critical to assess the potential impact of new interventions, such as vaccination campaigns implemented alone or in combination with vector control interventions [33,34], both prospectively and retrospectively. Across Africa, only 17 seroprevalence surveys have been conducted to date [3] and, as such, the African region remains a continent with limited knowledge of the historical circulation and seroprevalence of dengue [25]. Serological surveys are, therefore, invaluable surveillance tools to help understand and estimate dengue transmission intensity and its heterogeneity across the continent [35].

We propose a simulation-based method to guide secondary testing of existing serosurveys for other viruses. Retargeting existing serosurveys has been previously explored, for example by Carcelen et al. [36], who piggybacked a HIV national serosurvey performed in Zambia to test for anti-measles and anti-rubella virus IgG antibodies, using almost 10,000 residual sera. While testing existing samples may not be suitable in all circumstances, there are several advantages to secondary testing of recent serosurveys targeting a different virus, particularly those run during the COVID-19 pandemic. For instance, using existing samples from previously well-designed serosurveys reduces costs, resources and logistics, thus providing new opportunities for serosurveillance across multiple locations with a fixed budget.

This approach is particularly important, given the limited resources available for public health initiatives in many parts of the world, and our study contributes to the development of new strategies to make best use of available resources and improve our understanding of viral seroprevalence.

One limitation of the simulation approach adopted in this study is the use of imputed FOI estimates from the first global FOI map for dengue developed by Cattarino et al. [3] which, despite the accurate calibration, was be validated on a limited number of FOI estimates from the African region. In the absence of pre-existing information that can be used to inform the assumed FOI estimates, one possible strategy to overcome the intrinsic uncertainty associated with model estimates would be to implement serological surveys in two steps, e.g., by (1) testing half of the sample sizes identified in the simulation study according to the optimal age-distribution based on initial calculations, which can then be used to update the simulation study and inform the age-distribution, before (2) testing the other half of the samples (Fig 4). Another limitation of the method we developed is the empirical choice of the tolerance used to determine sample sizes – such as the 15% difference from the upper bound of the 95% CrI of the baseline scenario – which can be tweaked depending on the targeted accuracy and available budget and resources. Nonetheless, the methodological framework developed in this study provides a rationale that can be applied to secondary testing of previous serosurveys against different infectious diseases at variable spatial resolution, while optimizing the accuracy of the FOI estimates and the use of resources, also in locations where there is limited previous information on the historical circulation and transmission intensity of a pathogen.

**Fig 4.**
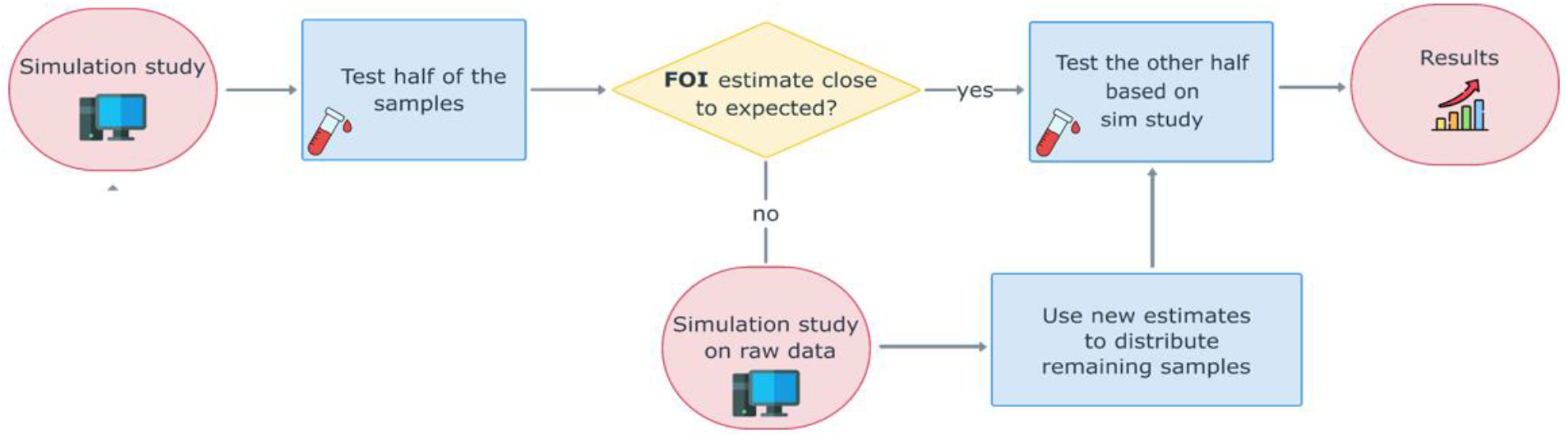
Conceptual description of the workflow for the implementation of serological surveys in the absence of previous information (such as serosurvey or case-notification data) on the FOI. Illustration of the two-step process proposed to identify the sample sizes and optimize their age-distribution when the study is conducted in locations with little or no prior information on dengue FOI.

## Conclusion

Age-stratified serological surveys are an ideal surveillance tool for reconstructing the age-dependent immunity profile of a population against one or more circulating viruses. Because the implementation of new serological surveys requires significant financial and human resources, testing stored blood samples (e.g., from previous serological surveys) can be an alternative strategy to estimate the FOI. In this work, we developed a method to inform the number of samples and age-distribution to test when using existing samples. In the presence of large uncertainties around the accuracy of the FOI estimates, we propose implementing the serological survey in two-steps, including an interim analysis of the results obtained, for instance on half of the number of samples identified, with the simulation-based analysis. The method developed in this paper can be used when testing existing blood samples for antibodies against different diseases in multiple settings, including in locations with limited prior information on the historical circulation of the pathogen of interest.

## Data Availability

All data and code produced in the present study are available in the supplementary material.

## Funding

AV thanks the Foundation Blanceflor Boncompagni Ludovisi (Sweaden) for funding her PhD visiting period at Imperial College London.

CM, CAD and ID acknowledge funding from the Drugs for Neglected Diseases initiative and the MRC Centre for Global Infectious Disease Analysis (reference MR/R015600/1), jointly funded by the UK Medical Research Council (MRC) and the UK Foreign, Commonwealth Development Office (FCDO), under the MRC/FCDO Concordat agreement and is also part of the EDCTP2 programme supported by the European Union.

ID acknowledges research funding from a Sir Henry Dale Fellowship funded by the Royal Society and Wellcome Trust (grant 213494/Z/18/Z).

CAD thanks the UK National Institute for Health and Care Research Health Protection Research Unit (NIHR HPRU) in Emerging and Zoonotic Infections in partnership with Public Health England (PHE) for funding (grant HPRU200907).

DNDi thanks the French Development Agency (AFD), France ; Médecins Sans Frontières International; Swiss Agency for Development and Cooperation (SDC), Switzerland; UK aid, UK; for the financial support in this work. The findings and conclusions contained herein are those of the authors and do not necessarily reflect positions or policies of the aforementioned funding bodies.

## Contributions

Conceptualisation: ID, BP, IR, NM, CAD

Data curation: AV, ID

Formal analysis: AV, ID

Methodology: AV, ID

Software: AV, CMC, ID

Validation: AV, CMC

Visualization: AV, CMC, CAD, ID

Writing-Original Draft Preparation: AV, CMC, ID, CAD

Writing- review & editing: AV, CMC, ID, BP, IR, NM, JHA, AA, CO, NSS, EL, JM, CAD

Funding acquisition: ID, BP, IR, NM

Project administration: ID, BP, IR, NM, JHA,

Resources: BP, IR, NM, JHA, AA, CO, NSS, EL, JM

Investigation: JHA, AA, CO, NSS, EL, JM

Supervision: ID, CAD

## Data availability statement

All data and code are available as Supplementary material.

## Notes

### Competing Interest Statement

The authors have declared no competing interest.

### Author Declarations

Ethics committee of Kumasi Centre for Collaborative Research in Tropical Medicine (KCCR) gave ethical approval for this work.

