## Supplementary Information for "A simulation-based method to inform serosurvey designs for estimating dengue force of infection using existing blood samples"

#### Table of Contents

Figures ........ 2
Tables ........ 3

### Figures

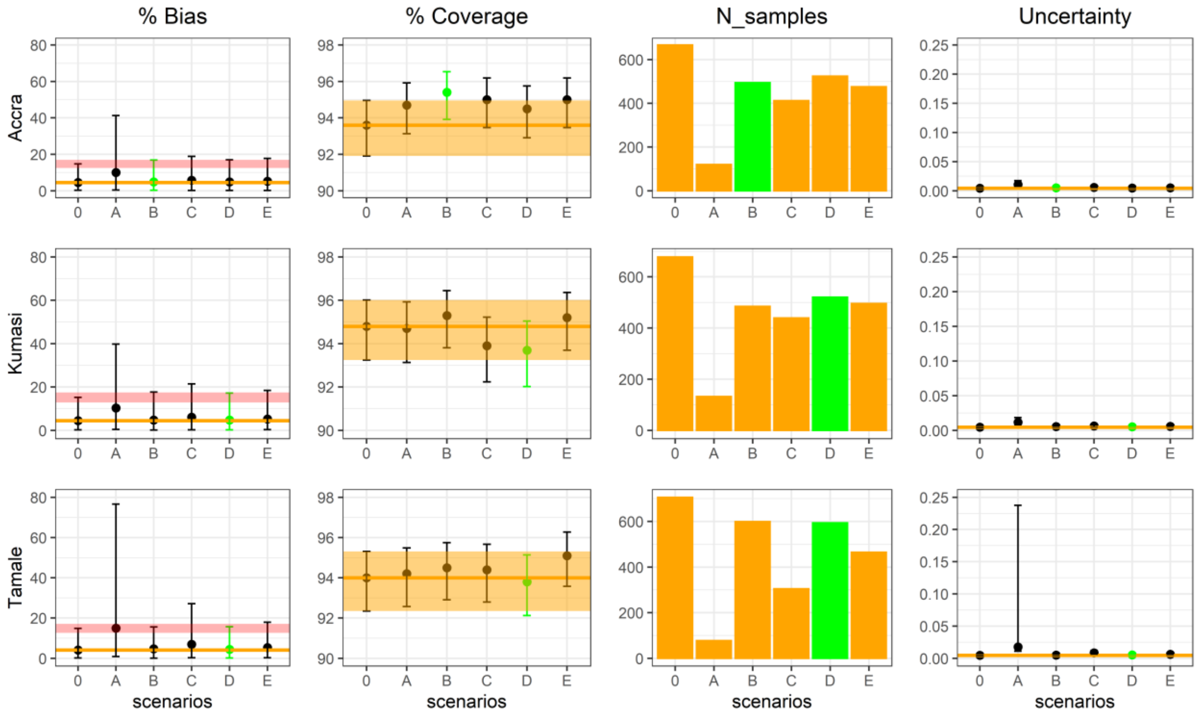


**Figure S1: Summary of the accuracy metrics obtained for Accra, Kumasi and Tamale with 10-year age category across scenarios.** The four columns represent the bias, coverage, number of tested samples and uncertainty obtained for each scenario. The scenario highlighted in green indicates the selected scenario. The median bias and uncertainty are reported with their 95% CrI (columns 1 and 4), while the median coverage is reported with its 95% exact binomial CI. The orange line represents the median (columns 1, 2 and 4) and the orange ribbon represents the 95% CrI of the baseline scenario 0 (columns 2 and 4). The pink ribbon in the first columns represents the 15% tolerance around the upper bound of the 95% CrI of scenario 0, which was used in the first step of the selection criterion.

### Tables

**Table S1** Sample sizes for 5-year age categorisation used in scenarios 0-E for Accra

| Age group 5 years | Scenario 0 | Scenario A | Scenario B | Scenario C | Scenario D | Scenario E |
|---|---|---|---|---|---|---|
| **10-14** | 20 | 20 | 20 | 16 | 20 | 10 |
| **15-19** | 57 | 20 | 57 | 16 | 57 | 28 |
| **20-24** | 81 | 20 | 81 | 16 | 81 | 40 |
| **25-29** | 90 | 20 | 90 | 16 | 90 | 45 |
| **30-34** | 69 | 20 | 69 | 16 | 69 | 34 |
| **35-39** | 64 | 20 | 64 | 16 | 64 | 32 |
| **40-44** | 44 | 20 | 10 | 44 | 22 | 44 |
| **45-49** | 66 | 20 | 10 | 66 | 33 | 66 |
| **50-54** | 45 | 20 | 10 | 45 | 22 | 45 |
| **55-59** | 38 | 20 | 10 | 38 | 19 | 38 |
| **60-64** | 31 | 20 | 10 | 31 | 16 | 31 |
| **65+** | 62 | 20 | 10 | 62 | 31 | 62 |
| **Total** | 667 | 240 | 441 | 382 | 524 | 475 |

**Table S2** Sample sizes for 5-year age categorisation used in scenarios 0-E for Kumasi.

| Age group 5 years | Scenario 0 | Scenario A | Scenario B | Scenario C | Scenario D | Scenario E |
|---|---|---|---|---|---|---|
| **10-14** | 22 | 22 | 22 | 17 | 22 | 11 |
| **15-19** | 58 | 22 | 58 | 17 | 58 | 29 |
| **20-24** | 88 | 22 | 88 | 17 | 88 | 44 |
| **25-29** | 82 | 22 | 82 | 17 | 82 | 41 |
| **30-34** | 65 | 22 | 65 | 17 | 65 | 32 |
| **35-39** | 50 | 22 | 50 | 17 | 50 | 25 |
| **40-44** | 61 | 22 | 11 | 61 | 30 | 61 |
| **45-49** | 65 | 22 | 11 | 65 | 32 | 65 |
| **50-54** | 51 | 22 | 11 | 51 | 26 | 51 |
| **55-59** | 34 | 22 | 11 | 34 | 17 | 34 |
| **60-64** | 43 | 22 | 11 | 43 | 22 | 43 |
| **65+** | 59 | 22 | 11 | 59 | 30 | 59 |
| **Total** | 678 | 264 | 431 | 415 | 522 | 495 |

**Table S3** Sample sizes for 5-year age categorisation used in scenarios 0-E for Tamale.

| Age group 5 years | Scenario 0 | Scenario A | Scenario B | Scenario C | Scenario D | Scenario E |
|---|---|---|---|---|---|---|
| **10-14** | 14 | 13 | 14 | 6 | 14 | 7 |
| **15-19** | 64 | 13 | 64 | 6 | 64 | 32 |
| **20-24** | 100 | 13 | 100 | 6 | 100 | 50 |
| **25-29** | 113 | 13 | 113 | 6 | 113 | 56 |
| **30-34** | 118 | 13 | 118 | 6 | 118 | 59 |
| **35-39** | 73 | 13 | 73 | 6 | 73 | 36 |
| **40-44** | 76 | 13 | 7 | 76 | 38 | 76 |
| **45-49** | 32 | 13 | 7 | 32 | 16 | 32 |
| **50-54** | 41 | 13 | 7 | 41 | 20 | 41 |
| **55-59** | 13 | 13 | 7 | 13 | 6 | 13 |
| **60-64** | 23 | 13 | 7 | 23 | 12 | 23 |
| **65+** | 39 | 13 | 7 | 39 | 20 | 39 |
| **Total** | 706 | 156 | 524 | 260 | 594 | 464 |

**Table S4** Sample sizes for 10-year age categorisation used in scenarios 0-E for Accra.

| Age group 10 years | Scenario 0 | Scenario A | Scenario B | Scenario C | Scenario D | Scenario F |
|---|---|---|---|---|---|---|
| **10-19** | 77 | 20 | 77 | 42 | 77 | 38 |
| **20-29** | 171 | 20 | 171 | 42 | 171 | 86 |
| **30-39** | 133 | 20 | 133 | 42 | 133 | 66 |
| **40-49** | 110 | 20 | 38 | 110 | 55 | 110 |
| **50-59** | 83 | 20 | 38 | 83 | 42 | 83 |
| **≥60** | 93 | 20 | 38 | 93 | 46 | 93 |
| **Total** | 667 | 120 | 495 | 412 | 524 | 476 |

**Table S5** Sample sizes for 10-year age categorisation used in scenarios 0-E for Kumasi.

| Age group 10 years | Scenario 0 | Scenario A | Scenario B | Scenario C | Scenario D | Scenario E |
|---|---|---|---|---|---|---|
| **10-19** | 80 | 22 | 80 | 42 | 80 | 40 |
| **20-29** | 170 | 22 | 170 | 42 | 170 | 85 |
| **30-39** | 115 | 22 | 115 | 42 | 115 | 58 |
| **40-49** | 126 | 22 | 40 | 126 | 63 | 126 |
| **50-59** | 85 | 22 | 40 | 85 | 42 | 85 |
| **≥60** | 102 | 22 | 40 | 102 | 51 | 102 |
| **Total** | 678 | 132 | 485 | 439 | 521 | 496 |

**Table S6** Sample sizes for 10-year age categorisation used in scenarios 0-E for Tamale.

| Age group 10 years | Scenario 0 | Scenario A | Scenario B | Scenario C | Scenario D | Scenario E |
|---|---|---|---|---|---|---|
| **10-19** | 78 | 13 | 78 | 27 | 78 | 39 |
| **20-29** | 213 | 13 | 213 | 27 | 213 | 106 |
| **30-39** | 191 | 13 | 191 | 27 | 191 | 96 |
| **40-49** | 108 | 13 | 39 | 108 | 54 | 108 |
| **50-59** | 54 | 13 | 39 | 54 | 27 | 54 |
| **≥60** | 62 | 13 | 39 | 62 | 31 | 62 |
| **Total** | 706 | 78 | 599 | 305 | 594 | 465 |

**Table S7** Age-specific samples sizes for the 5-years age categorisation with the selected scenario.

| Age group 5-years | Accra | Kumasi | Tamale |
|---|---|---|---|
| **Scenario** | **B** | **D** | **B** |
| **10-14** | 20 | 22 | 14 |
| **15-19** | 57 | 58 | 64 |
| **20-24** | 81 | 88 | 100 |
| **25-29** | 90 | 82 | 113 |
| **30-34** | 69 | 65 | 118 |
| **35-39** | 64 | 50 | 73 |
| **40-44** | 10 | 30 | 7 |
| **45-49** | 10 | 32 | 7 |
| **50-54** | 10 | 26 | 7 |
| **55-59** | 10 | 17 | 7 |

| | | | |
|---|---|---|---|
| **60-64** | 10 | 22 | 7 |
| **65+** | 10 | 30 | 7 |
| **Total** | 441 | 522 | 524 |

**Table S8** Age-specific samples sizes for the 10-years age categorisation with the selected scenario.

| **Age group 10-years** | **Accra** | **Kumasi** | **Tamale** |
|---|---|---|---|
| **Scenario** | **B** | **D** | **D** |
| **10-19** | 77 | 80 | 78 |
| **20-29** | 171 | 170 | 213 |
| **30-39** | 133 | 115 | 191 |
| **40-49** | 38 | 63 | 54 |
| **50-59** | 38 | 42 | 27 |
| **≥60** | 38 | 51 | 31 |
| **Total** | 495 | 521 | 594 |

**Table S9** City-specific FOI estimates from Cattarino et al [3].

| | **Accra** | **Kumasi** | **Tamale** |
|---|---|---|---|
| **FOI** | 0.017 | 0.019 | 0.020 |
